# The role of *KPNA2* mutations in breast cancer prognosis: A survey of publicly available databases

**DOI:** 10.1101/2021.11.10.21266193

**Authors:** Layla Alnoumas, Lisa van Den Driest, Alison Lannigan, Caroline H Johnson, Nicholas JW Rattray, Zahra Rattray

## Abstract

Breast cancer, comprising of several sub-phenotypes, is a leading cause of female cancer-related mortality in the UK and accounts for 15% of all cancer cases. Chemoresistant sub phenotypes of breast cancer remain a particular challenge. However, the rapidly-growing availability of clinical datasets, presents the scope to underpin a data driven precision medicine-based approach exploring new targets for diagnostic and therapeutic interventions. We report a survey of several publicly available databases probing the expression and prognostic role of Karyopherin-2 alpha (KPNA2) in breast cancer prognosis. Aberrant *KPNA2* overexpression is directly correlated with aggressive tumour phenotypes and poor patient survival outcomes. We examined the existing information available on a range of commonly occurring mutations of *KPNA2* and their correlation with patient survival.

Our analysis of clinical gene expression datasets show that *KPNA2* is frequently amplified in breast cancer, with differences in expression levels observed as a function of patient age and clinicopathologic parameters. We also found that aberrant *KPNA2* overexpression is directly correlated with poor patient prognosis, warranting further investigation of KPNA2 as an actionable target for patient stratification or the design of novel chemotherapy agents.

In the era of big data, the wealth of datasets available in the public domain can be used to underpin proof of concept studies evaluating the biomolecular pathways implicated in chemotherapy resistance in breast cancer.

## Introduction

Breast cancer is the most commonly-diagnosed, and leading cause of cancer-related mortality worldwide among women with an estimate of 2.3 million new cases in 2020 [1, 2]. Breast cancer represents a heterogeneous group of diseases classified across several sub-phenotypes according to their anatomical location and gene expression profile.

Despite significant advancements in developing new treatments for breast cancer, the incidence of breast cancer in women continues to rise proportionally with age, posing a significant global public health challenge [3]. Current standard of care in breast cancer treatment involves surgery, radiotherapy, endocrine-based therapies, chemotherapies or biologicals, or a combination of these therapeutic interventions. From a diagnostic perspective, mammography remains one of the main approaches for detecting breast cancer. However, patients are often diagnosed during later stages of breast cancer with the potential to adversely impact patient clinical prognosis and outcomes. Therefore, the recent years have seen a significant growth in novel surrogate biomarker research for diagnostic, prognostic and therapeutic interventions. Current routine stratification for breast cancer treatment is based on the hormonal status (oestrogen, progesterone and human epidermal growth receptor-2) or more recently, genetic biomolecular signatures classifying breast cancers according to intrinsic subtypes (e.g. basal and luminal A and B) [4].

Karyopherin alpha 2 (KPNA2), a member of the Karyopherin family and an adaptor protein, is a component of the nuclear import pathway machinery involved in the nucleocytoplasmic transport of molecules involved in cell division, transcription, and DNA repair. Aberrant amplification of *KPNA2* expression in cancer has been implicated in the pathogenic mislocalization of substrate proteins, resulting in tumorigenesis and conferring an aggressive sub-phenotype [5]. KPNA2 over-expression has been correlated with poor patient outcomes in a number of malignancies including glioblastoma [6], colon [7], hepatocellular carcinoma [8], ovarian [9] and breast [10-12] cancers. In breast cancer, *KPNA2* expression is correlated with a lower abundance of DNA repair proteins including CHK1, UBC9, PIAS1, BRCA1, RAD51 and γH2AX in cell nuclei [12]. Moreover, the incidence of KPNA2 overexpression is correlated with oestrogen receptor-negative (ER-) status [12, 13].

With increasing reports of KPNA2 involvement in several cancer types [6, 7, 9, 14] and significant advancements in precision medicine technologies, coupled to extensive biobanking and electronic curation of patient metadata, the scope exists to interrogate the correlation between *KPNA2* expression, breast cancer phenotype and patient prognosis. Dysregulation of mRNA expression levels of *KPNA2* in human breast cancer and its association with breast cancer prognosis has not yet been investigated. In this study, we report the use of a range of bioinformatics tools to investigate the roles of KPNA2 in human breast cancer. We analyzed the mRNA expression patterns and mutations of *KPNA2* in patients with breast cancer from the vast number of gene expression data available within the public domain, to identify expression patterns and the potential prognostic value of KPNA2 in human breast cancer.

## Materials and Methods

### Data retrieval

**cBioPortal** (https://www.cbioportal.org/) is an open access resource for cancer genomics that was originally developed by Memorial Sloan Kettering Cancer Center [15]. In this study cBioPortal was used to query the incidence and types of *KPNA2* mutations occurring in breast cancer as a function of tumour clinicopathologic parameters.

**COSMIC** (Catalogue of Somatic Mutations in Cancer (www.sanger.ac.uk)) is a tool for studying the influence of somatic mutation in all cancers and assessing druggability of targets incorporation with chEMBL, which is maintained by the European Molecular Biology Laboratory. Using this resource, we identified over 500 *KPNA2*-related mutations, specifying the amino acid point mutation position and mutation type, whether missense or insertion.

**Oncomine** (https://www.oncomine.org/resource/login.html) Analysis of *KPNA2* mRNA expression patterns was conducted using the following parameter selections: Gene-*KPNA2*, differential analysis-cancer vs. normal analysis, cancer type-breast cancer; and data type-mRNA. A two-fold change, a P-value corresponding to 1E-4 and a top 10% gene rank were selected as thresholds for this analysis. The same parameters were applied to the analysis of gene co-expression analyses. All statistical analyses and parameters were directly exported from Oncomine.

**PrognoScan** (http://dna00.bio.kyutech.ac.jp/PrognoScan/index.html) [16] is a resource for performing meta-analysis of the prognostic role of mutations occurring in cancer through incorporating gene expression studies from multiple sources such as the Gene Expression Omnibus (GEO - www.ncbi.nlm.nih.gov/gds) and reports from individual labs [17]. PrognoScan combines expression data with clinical outcomes, which enables the evaluation of potential biomarkers and their role in cancer prognosis. In this study PrognoScan was used to assess the correlation between *KPNA2* mRNA expression levels and patient prognostic endpoints for breast cancer. Output generated and exported from PrognoScan include P-values (Cox), hazard ratios and confidence intervals across breast cancer datasets available. Data available for the 201088_at *KPNA2* reporter was selected for the generation of Forest plots.

**Kaplan-Meier Plotter (KMplot)** (http://kmplot.com/analysis/index.php?p=service) [18] uses gene expression data from GEO datasets and through integration will clinical data, generates Kaplan-Meier plots across multiple prognostic outcomes. Using this tool it is possible to restrict the selection to patients with specific breast cancer sub-phenotypes, enabling the selection of inclusion and exclusion criteria. For the purposes of this study, the prognostic value of KPNA2 was studied across all breast cancer types, and as a function of each intrinsic molecular subtype (St. Gallen definitions were used) [19]. For all survival analyses, the auto select best cut-off was used to display the P-value (log-rank) and false-discovery rate (FDR) for each plot and the probe ID (201088_at) of *KPNA2* reporter was selected for all searches.

**Breast Cancer Gene-Expression Miner v4.6 [20]** (http://bcgenex.ico.unicancer.fr/BC-GEM/GEM-Accueil.php?js=1) is a breast cancer statistical mining tool providing information on gene expression and prognostic implications of gene expression profiles in breast cancer. Moreover, the correlation between multiple genes, and their association with breast cancer can be elucidated using this tool [21]. Briefly, *KPNA2* expression patterns in all breast cancers were examined (RNA-seq, all platforms) and endpoint events (overall survival, disease-free survival) classified according to sub-phenotypes. Gene ontology and exhaustive gene correlations were also studied across all breast cancer groups as a function of intrinsic molecular subtype and hormone receptor expression profile.

### Statistical Analysis

Comparisons of *KPNA2* mRNA expression levels performed between breast cancer and healthy breast tissue (fold-change) was performed in Oncomine using a t-test. For comparisons between breast cancer patient subsets in Geneminer, a Welch test was used to compare differences in KPNA2 mRNA expression. To analyze the prognostic value of *KPNA2* using Kaplan-Meier plot (KMPlot), P-values from log-rank analysis were used to compare prognostic endpoints between patient cohorts using in-built algorithms on the webpage. Prognostic data obtained from PrognoScan was selected according to the calculated Cox P-values and corresponding Hazard ratios (95% confidence interval) for various endpoints (overall survival, disease-free survival, disease-free metastatic survival, and relapse-free survival) that were subsequently plotted and visualized with a Forest plot. Unless otherwise stated, a P<0.05 was deemed as statistically significant for all comparisons.

## Results

### KPNA2 mutations in breast cancer

Gene alterations impacting *KPNA2* in breast cancer were analyzed using cBioPortal and COSMIC databases. Querying a combined total of 4,065 samples across five studies in cBioportal, the frequency of *KPNA2* gene alterations differed across each study queried (Table 1). The percentage of samples with somatic mutations in *KPNA2* were 0.2% of the *KPNA2*-related duplicate mutations, corresponding to 8 missense substitutions and one in-frame deletion in patients with multiple samples (see supplementary information). Amplification of *KPNA2* expression was the most frequently observed alteration across all As a validation step, patterns of *KPNA2* expression were studied across 39,619 cancer samples in COSMIC. These analyses revealed that 341 out of 2,612 breast cancer samples contained seven KPNA2 amino acid changes characterized as missense mutations. Six out of the seven mutations identified in COSMIC were identical to those found in cBioPortal. In the case of COSMIC, no deletion mutations were found in the *KPNA2* sequence, but an additional missense mutation (A364V) was present (**Table 2**).

**Table 1.** A summary of breast cancer studies located from cBioportal and corresponding frequency of mutations

| Study | Percent of total cases<br>(Proportion of cases<br>available in dataset) | Frequency |
| --- | --- | --- |
| The Metastatic Breast<br>Cancer Project [22] | 15.6%(37/237) | Mutation (1.27%, n=3)<br>Amplification (14.4%, n=34) |
| Metastatic Breast<br>Cancer (INSERM)[23] | 7.87%(17/216) | Amplification (7.87%, n=17) |
| Breast Cancer<br>(Metabric)[24-26] | 7.55%(164/2173) | Amplification (7.5%, n=163)<br>Deep Deletion (0.05%, n=1) |
| TCGA Pan-cancer Atlas<br>[27] | 6.46%(70/1084) | Mutation (0.55%, n=6)<br>Amplification (5.9%, n=64) |

studies examined.

**Table 2.** Corresponding particulars of mutations occurring in *KPNA2* and their frequency located in cBioportal. ***TCGA:*** The Cancer Genome Atlas Program Pan-Cancer Atlas [27] and the ***MBC:*** Metastatic Breast Cancer Project [22].

| Protein change | Mutation type | No. of<br>mutations | Variant<br>frequency | Original study |
| --- | --- | --- | --- | --- |
| **R366H** | Missense | 1250 | 0.23 | TCGA |
| **N375S** | Missense | 972 | 0.24 | TCGA |
| **L382F** | Missense | 832 | 0.07 | TCGA |
| **S24N** | Missense | 123 | 0.06 | MBC |
| **V507A** | Missense | 30 | 0.28 | MBC |
| **D79N** | Missense | 107 | 0.23 | TCGA |
| **Q329del** | Deletion | 36 | 0.30 | TCGA |
| **R29C** | Missense | 35 | N/A | TCGA |

Next, we assessed the correlation between amplification of *KPNA2* mRNA expression levels in breast cancer tumours compared to matched healthy breast tissue using Oncomine. Findings from these comparisons across intrinsic molecular subtype and corresponding fold-changes are presented in **Table 3**.

**Table 3.** *KPNA2* expression is frequently amplified in breast cancer in comparison to healthy tissue. Breast cancer subtypes and corresponding fold-changes in *KPNA2* expression relative to adjacent breast tissue for datasets located in Oncomine. P-values are directly exported from Oncomine and are obtained from a two-sample t-test.

| Breast Cancer Subtype | Fold-change | P-value | Patient numbers | Overexpression Gene Rank | Study Ref |
| --- | --- | --- | --- | --- | --- |
| Ductal Breast Carcinoma | 5.3 | 6.62E-18 | 40 | Top 1% | Richardson [21] |
| Male Breast Carcinoma | 4.7 | 1.84E-31 | 3 | Top 1% | TCGA [27] |
| Invasive Ductal Breast Carcinoma | 3.2 | 5.54E-47 | 389 | Top 1% | TCGA [27] |
| Medullary Breast Carcinoma | 2.6 | 1.23E-8 | 32 | Top 7% | Curtis [23] |
| Invasive Ductal Breast Carcinoma | 2.2 | 9.54E-81 | 1,556 | Top 3% | Curtis [23] |
| Invasive Lobular Breast Carcinoma | 2.1 | 1.58E-11 | 36 | Top 4% | TCGA [27] |
| Ductal Breast Carcinoma *In Situ* | 2.1 | 8.67E-6 | 11 | Top 1% | Ma [22] |
| Breast Carcinoma | 2.1 | 4.00E-5 | 14 | Top 5% | Curtis [23] |
| Invasive Breast Carcinoma | 2.1 | 1.99E-5 | 21 | Top 7% | Curtis [23] |

Our analysis of fold-change data show that within breast cancer datasets available on Oncomine, *KPNA2* frequently was ranked in the top 7% of genes altered in breast cancer with significant fold-changes observed across all studies relative to adjacent breast cancer tissue. Across all breast cancer subtypes examined, at least a positive two-fold increase (with a corresponding P-value <0.05) was observed in *KPNA2* mRNA expression levels between healthy and breast cancer tissue, indicating *KPNA2* overexpression across various breast cancer types.

Next, we performed a search of the patterns of *KPNA*2 mRNA expression in breast cancer using Oncomine, cBioPortal and Geneminer toolsets. Analysis of the datasets available on these resources indicated differential *KPNA2* expression levels as a function of clinicopathological parameters (**Fig. 1**).

**Figure 1.**
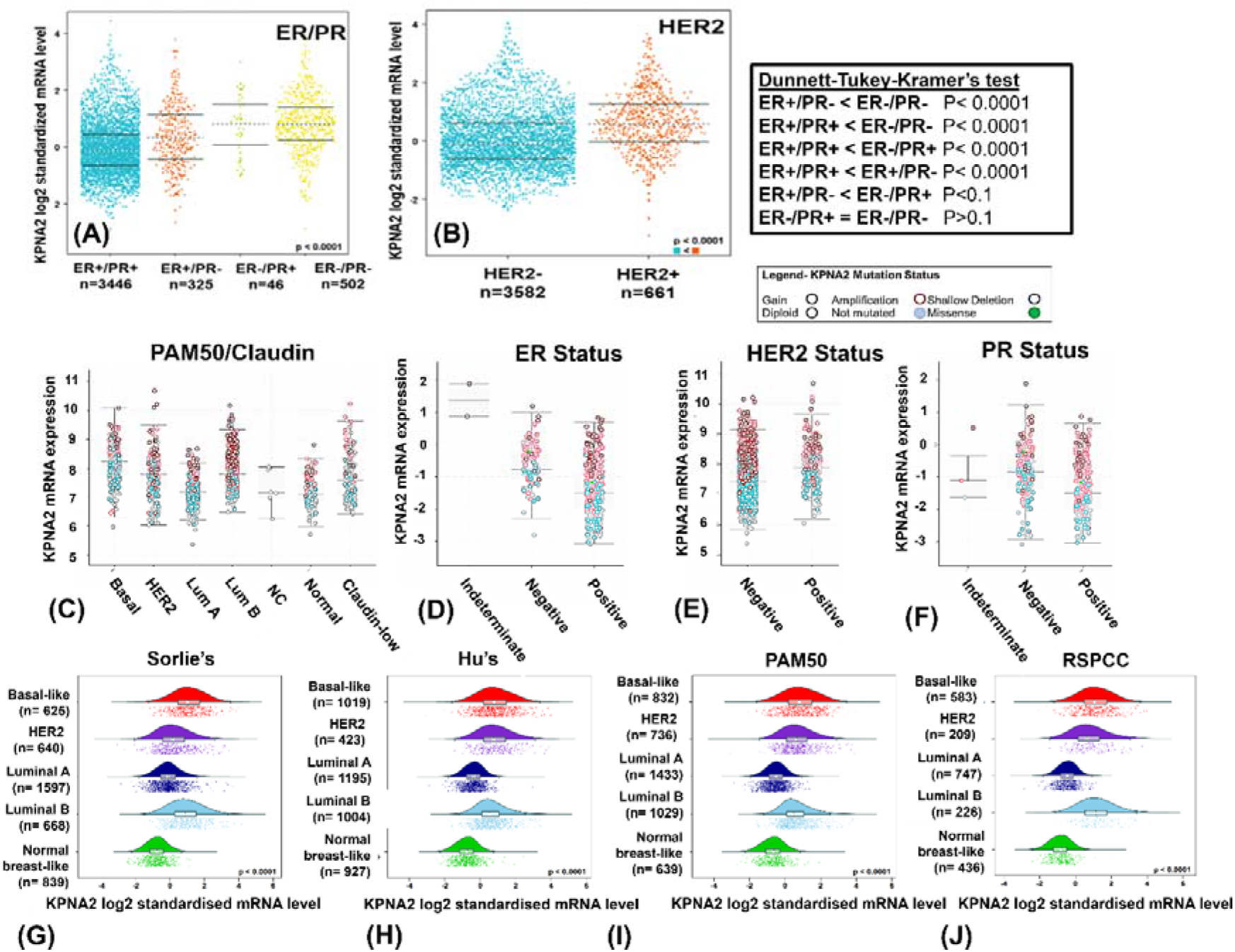
*KPNA2* mRNA expression varies as a function of breast cancer clinicopathologic parameters. Bee swarm plots of *KPNA2* mRNA expression levels as a function of combined oestrogen (ER) and progesterone (PR) receptor status (A) and HER2 receptor status (B) across breast cancer studies obtained from Geneminer. Boxplots of *KPNA2* mRNA expression levels as a function of PAM50 molecular subtype status (C), oestrogen (D), HER2 (E), and progesterone (F) receptor status for data located on cBioPortal. Corresponding *KPNA2* mRNA levels according to Sorlie’s (G), Hu’s (H), PAM50 (I), and RSPCC (J) intrinsic molecular subtypes located in Geneminer.

Analysis of *KPNA2* expression level patterns across multiple toolsets shows a varied *KPNA2* expression and mutational profile as a function of clinicopathological parameters. The incidence of *KPNA2* genetic alterations occurred more frequently in patients with positive ER status (**Fig. 1F**), whereas higher *KPNA2* mRNA levels appeared in patients with negative hormone receptor status (**Fig. 1 A-B**). Relative to normal breast-like tissue, mRNA expression levels of *KPNA2* are significantly elevated across all molecular subtypes. Across Geneminer and Oncomine databases, *KPNA2* amplification occurred most frequently in patients aged <40 years in comparison to post-menopausal patients (see supplementary information, Welch’s P<0.0001, GeneMiner). We also compared *KPNA2* expression profiles across different breast cancer subtypes that included carcinoma, invasive ductal carcinoma and adenocarcinoma. *KPNA2* amplification occurred in patients with invasive ductal carcinoma and was more frequently observed in patients with oestrogen-receptor negative breast cancer. Pairwise comparisons of the relative *KPNA2* mRNA expression levels were performed in Geneminer according to tumour intrinsic molecular subtype. Corresponding readout indicates differential *KPNA2* expression patterns across the sub-phenotypic classifications, with normal breast-like tumours consistently exhibiting (statistically significant, P<0.0001) lower *KPNA2* expression levels in comparison to other molecular sub-phenotypes.

### Aberrant *KPNA2* expression is associated with poor breast cancer prognosis

The prognostic value of *KPNA2* in breast cancer was examined using PrognoScan and KMPlot. In PrognoScan, 25 Gene Expression Omnibus (GEO) datasets were located in total, which were divided across five categories of 10 distant metastasis-free survival (DMFS), 2 Disease-free survival (DFS), 2 Disease-specific survival (DSS), 8 Relapse-free survival (RFS), and 3 overall survival (OS). Data presented in the Forest plot consistently demonstrate a negative correlation between *KPNA2* overexpression and patient survival (**Fig. 2**).

**Figure 2.**
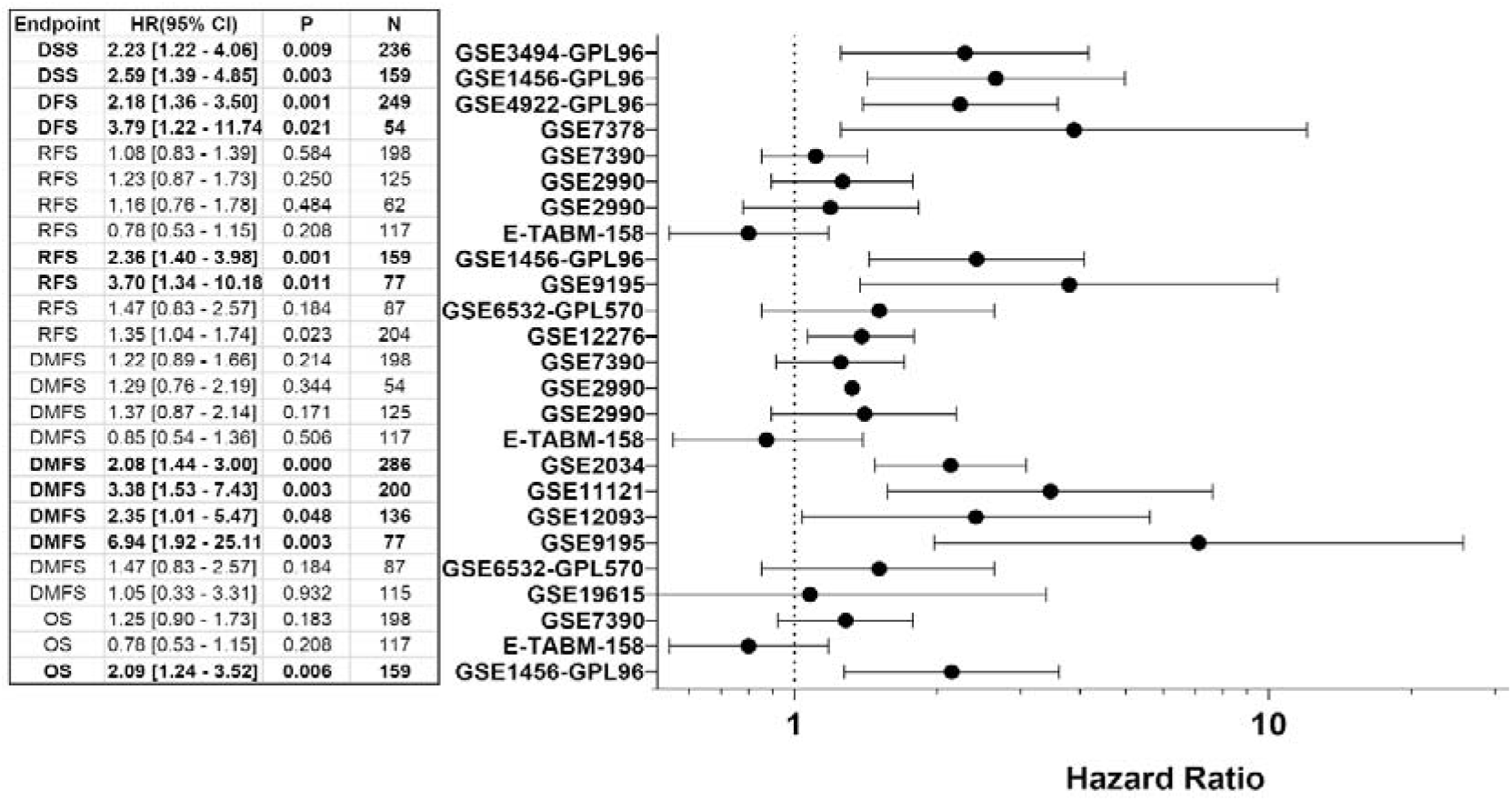
*KPNA2* overexpression is associated with poor prognostic outcomes. *Forest plot representing the association between KPNA2 expression and prognostic outcomes for studies using the* 201088_at *KPNA2* reporter.

The number of breast cancer dataset entries extracted from PrognoScan across all *KPNA2* reporters were 57 studies in total. These were further categorized into one of five categories including relapse-free survival (RFS- 18), disease-free survival (DFS- 5), disease-specific survival (DSS- 6), overall survival (OS- 8), and distant metastasis-free survival (DMFS- 19). The forest plot (**Fig. 2**) demonstrates a direct correlation between amplification of *KPNA2* expression and a poor prognosis across all endpoints.

The prognostic value of *KPNA2* overexpression across various breast cancer intrinsic molecular subtypes was studied, that included basal-like, luminal A, luminal B and HER2^+^ malignancies. As shown in **Fig. 3**, elevated *KPNA2* mRNA expression across all breast cancer types was associated with poorer OS (HR 1.68, CI 95% 1.35-2.08-, P= 2.6E-6, **Fig.3A**), RFS (HR 1.58, CI 95% 1.42-1.76, P<1E-16, **Fig.3B**), DMFS (HR 1.73, CI 95% 1.42-2.1, P= 3.9E-8, **Fig.3C**) and had no statistically significant impact on PPS (HR 1.71, CI 95% 1.32-2.22, P= 3.8E-5, **Fig.3D**).

**Figure 3.**
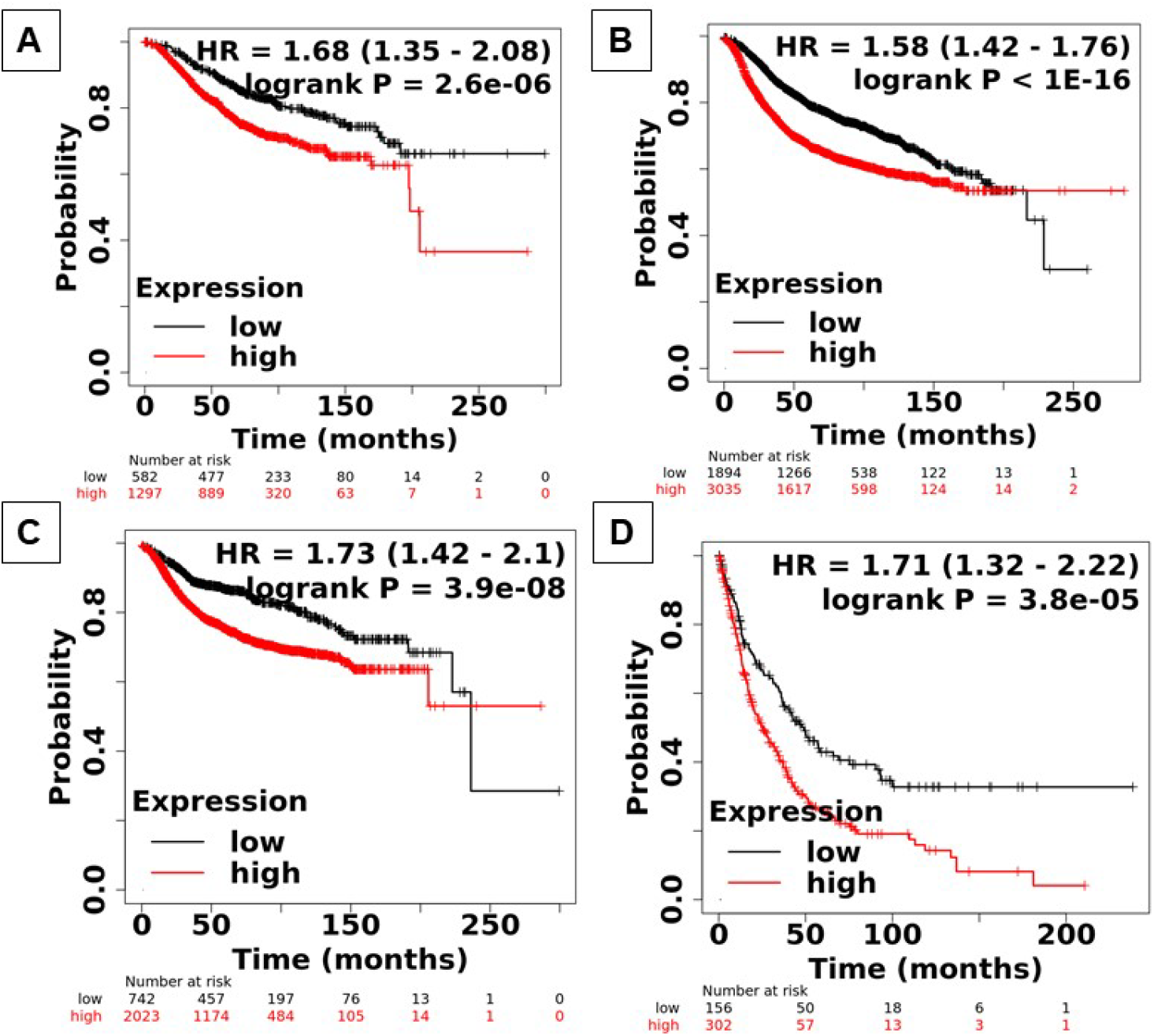
The prognostic value of *KPNA2* mRNA expression using Kaplan-Meier plotter (KMPlot) across all breast cancer types. Corresponding HRs for OS (A), RFS (B), DMFS (C), and PPS (D) survival endpoints across all breast cancer types. HR: Hazard ratio, BC: Breast Cancer, OS: Overall Survival, RFS: Relapse-free Survival, DMFS: Disease-Metastatic Free Progression Survival, and PPS: Post-Progression Survival.

Next, we examined the prognostic value of *KPNA2* mRNA expression across intrinsic molecular sub-phenotypes. From the datasets examined, elevated *KPNA2* mRNA levels had no significant overall prognostic impact on patients with basal carcinomas, Luminal B (except for RFS-HR 1.35, CI 95% 1.09-1.68, P= 0.0056, **Fig. 3N**) and HER2+ breast cancers. However, in the case of the Luminal A sub-phenotype, elevated *KPNA2* mRNA levels were associated with poor OS (HR 2.03, CI 95% 1.46-2.84-, P=2.2E-5, **Fig. 3I**), RFS (HR 1.73, CI 95% 1.46-2.04, P=9.6E-11, **Fig. 3J**), DMFS (HR 1.96, CI 95% 1.46-2.62, P= 4.1E-6, **Fig. 3K**) and PPS (HR 2.18, CI 95% 1.5-3.18, P=3.3E-5, **Fig. 3L**) outcomes. Overall, these findings show that *KPNA2* overexpression in breast cancer leads to poor patient survival outcomes across multiple endpoints, demonstrating the prognostic value of *KPNA2* as a potential biomarker and actionable target.

### Co-expression patterns of *KPNA2* mRNA in breast cancer

To identify the pathways impacted by aberrant KPNA2 activity, we examined the correlation in gene expression patterns between *KPNA2* and other genes using Oncomine. The top positive and negatively correlated genes with *KPNA2* are shown in **Fig. 4**. The Richardson Breast 2 study was selected to study gene co-expression patterns (P-value: 0.001, Fold change:2, Gene rank: 10%), with 186 located genes upregulated genes in ductal breast carcinoma.

**Figure 4.**
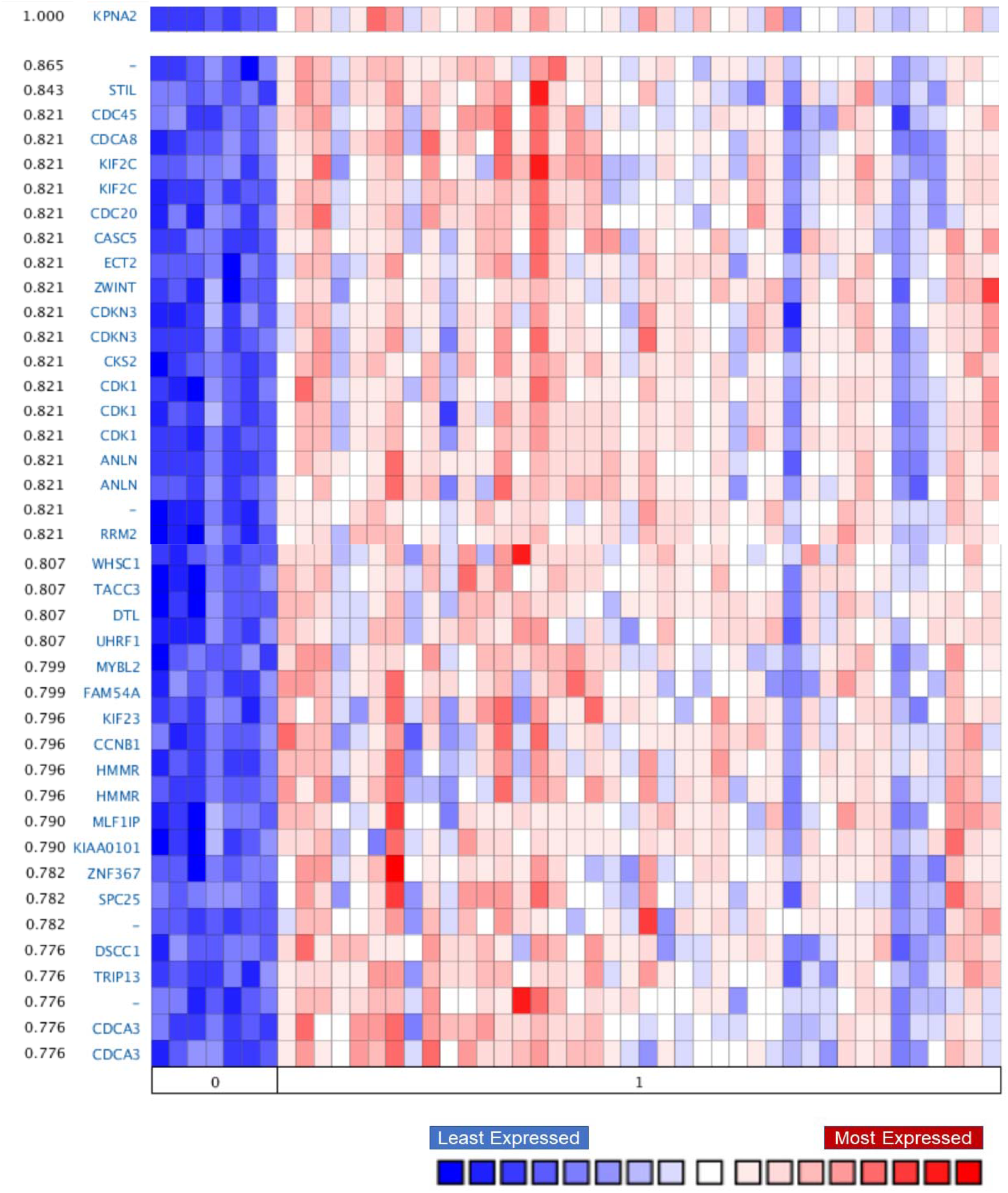
Heatmap of genes co-expressed with *KPNA2* in healthy breast tissue (0) and ductal breast carcinoma (1). Selected parameters from Oncomine included a fold-change of 3, a P-value of 0.001, and gene rank within the top 10%). (source: Richardson Breast Study 2, N=47 samples, and 19,574 measured genes) [28].

As shown in **Fig. 4**, genes most frequently co-expressed with *KPNA2* in ductal breast carcinoma were found to be least expressed in healthy breast tissue.

Taken together, our findings from analyses of *KPNA2* expression levels, mutational signature, impact on prognostic endpoints and co-expression patterns evidence that *KPNA2* is implicated in cancer progression and prognosis.

## Discussion

In the present study we examined the expression patterns of *KPNA2* and its prognostic significance in breast cancer as a function of clinicopathologic parameters using online bioinformatics databases. To-date, datasets from the genomic and transcriptomic-based analyses of breast cancer tumour biopsies and their corresponding metadata have been curated and deposited across multiple databases for public access as a precision medicine tool [20, 29]. To our knowledge, a comprehensive analysis of clinical datasets interrogating the frequency and patterns of *KPNA2* gene alterations as a function of tumour clinicopathologic parameters has not previously been attempted.

The dysregulation and aberrant function of Karyopherin activity has previously been correlated with tumour aggressiveness and poor patient prognosis across multiple cancer types. KPNA2, a member of the karyopherin family, is involved in the nucleocytoplasmic transport of a range of key cellular factors including DNA repair, transcription, and cell division factors [30]. Previous work has shown a direct correlation between *KPNA2* overexpression and poor patient prognosis across a range of cancer types, including glioblastoma, colorectal and ovarian cancer [6, 7, 9]. Despite the involvement of the Karyopherin family in breast cancer prognosis and tumorigenesis, the distinct role of KPNA2 in breast cancer outcomes and its expression patterns within breast tumour subtypes requires further investigation.

We used datasets available from online resources to analyse the frequency of genetic alterations occurring in *KPNA2* mRNA expression levels across breast cancer intrinsic molecular subtypes (Geneminer, cBioPortal, COSMIC and Oncomine), examined patterns of *KPNA2* co-expression with other genes (Geneminer and Oncomine) and evaluated the prognostic implications of *KPNA2* mRNA overexpression in patients with breast cancer (Prognoscan and Kaplan-Meier Plotter).

Our analysis of patterns of *KPNA2* mutations in cBioPortal and COSMIC revealed that N375S, a *MET* mutation, occurs across a range of cancer types and is detected in 9% of advanced breast tumours. This tyrosine kinase mutation has previously been shown to be oncogenic and dysregulated in early-stage lung cancers [31]. R366H mutations are common in colon cancer and involves a defective phosphorylation pathway of Long interspersed nuclear elements (LINE-1), activating inflammatory immune responses that drive tumour development [32]. Our searches of the Geneminer and cBioPortal repositories (**Fig. 1**) consistently show that the most frequently-occurring *KPNA2* genetic alteration in breast cancer tumours is overexpression. Furthermore, our results demonstrate that patients with hormone receptor-negative (ER/PR) status are most likely to exhibit higher *KPNA2* mRNA expression levels, in comparison to patients with hormone receptor-positive breast cancers (P<0.0001, **Fig. 1**). These data were further confirmed with the inverse correlation between *KPNA2*, and oestrogen and progesterone receptor mRNA levels (Geneminer, supplementary information). The incidence of *KPNA2* amplification was also found to be higher in younger patients with breast cancer (supplemental information), suggesting its role in breast cancer progression in this age group. Furthermore, *KPNA2* mRNA expression levels were found to be significantly amplified in patients with invasive ductal carcinoma (**Fig. 1B**).

Our search of the Oncomine database showed that at the transcriptional level relative to matched healthy breast tissue, the expression of KPNA2 was significantly upregulated in invasive lobular breast carcinoma, ductal breast carcinoma *in situ*, and invasive breast carcinoma. In all searches performed, *KPNA2* was ranked in the top 7% of genes dysregulated in cancer across breast cancer subtypes located.

Functional assessment of *KPNA2* co-expression showed that KPNA2 mRNA overexpression is directly correlated with an enrichment in genes regulating the cell cycle. SCL-interrupting locus protein (STIL), previously identified in prostate cancer [33], is a G2 phase gene involved in cell growth and development. This oncogene also activates the cell cycle-dependent protein kinase 1 (CDK1) pathway. *CDK1*, also co-expressed with *KPNA2*, promotes G2/M cell cycle transition and has previously been reported in hepatocellular carcinomas [8]. Moreover, *KPNA2* overexpression in ovarian cancer was recently linked to KIF4F signalling upregulation accelerating tumour progression [34, 35].

ZW10 interacting kinetochore protein (ZWINT) and Epithelial cell transforming 2 (ECT), both mitotic checkpoint proteins, have been shown to contribute to poor prognosis across multiple cancer types including glioblastoma [36]. Though previous reports show an association between ZWINT overexpression and triple-negative breast cancers, the functional role of ZWINT and ECT in breast cancer remains largely unexplored [37]. The *ECT* gene has been implicated in the protein assembly in cell division [38], and its dysregulation in breast cancer remains poorly understood. Another gene directly co-expressed with *KPNA2* is the Cell division cycle 20 (*CDC20*), a late mitosis checkpoint mediator that predominantly occurs in hormone positive (ER+) breast tumours (58% (N=870), METABRIC study) [39]. Aberrant *CDC20* overexpression has previously been implicated in pan-cancer disease progression and poor patient prognosis.

Our evaluation of the prognostic role of *KPNA2*, showed that across multiple prognostic endpoints (OS, RFS, DMFS and PPS) from PrognoScan and KMPlot (**Fig. 3**), KPNA2 overexpression was associated with poor survival outcomes. Our findings are in agreement with a previous report indicating that KPNA2 overexpression can serve as a prognostic marker across multiple cancer types and is associated with malignant transformation and poor patient survival [40-42].

To-date a limited number of reports have studied the functional role of KPNA2 in patient response to standard of care treatments and breast cancer outcomes. Our investigation primarily focused on using existing databases to inform the future rationale for exploring the biomolecular and phenotypic role of KPNA2 in breast cancer. Our integrated analyses of existing datasets indicate that KPNA2 can serve as a prognostic biomarker in breast cancer, warranting further investigation of its biomolecular role in tumour aggressiveness. We identified the functional associations and prognostic significance of KPNA2 in breast cancer, which warrants its further investigation as a promising prognostic biomarker or druggable target.

## Conclusion

During the COVID-19 pandemic and with limitations in laboratory access clinical datasets freely available on databases have provided a tool for data mining and scoping new projects. Open access databases provide a useful toolbox for investigation the correlations between biomolecular drivers of cancer and prognostic outcomes. Here, we used outputs from such databases to explore the rationale for targeting KPNA2 as a novel druggable target. Our analyses of existing clinical datasets for expression and survival outcomes show that KPNA2 over-expression contributes to poor patient survival outcomes, further necessitating its investigation in future studies to increase the range of treatments available for distinct breast cancer subtypes.

## Data Availability

All data produced in the present study are available upon reasonable request to the authors.

## CRediT authorship contribution statement

**Layla Alnoumas:** Methodology, Data curation, Formal analysis, Investigation, Writing – review & editing. **Lisa van Den Driest**: Methodology, Investigation. **Zoe Apczynski:** Investigation. **Alison Lannigan**: Writing – review & editing. **Caroline Johnson:** Data Interpretation, Funding acquisition, Writing-review & editing. **Nicholas Rattray:** Supervision, Writing – review & editing. **Zahra Rattray:** Conceptualization, Methodology, Investigation, Supervision, Funding acquisition, Formal analysis, Writing – original draft, Writing – review & editing.

## Conflicts of Interest

The authors declare no known competing financial interests or personal relationships that could have influenced the work reported in this manuscript.

## Acknowledgments

We acknowledge funding awarded to Zahra Rattray from Tenovus Scotland, The Royal Society of Edinburgh Research Reboot, FRAME summer studentship for LVD internship, and Kuwait University for LA PhD scholarship funding. We acknowledge support to Caroline Johnson from the American Cancer Society Research Scholar Grant 134273-RSG-20-065-01-TBE and the US National Cancer Institute of the National Institutes of Health under Award Number K12CA215110. We dedicate this manuscript to all the patients with breast cancer who support ongoing efforts in developing new diagnostics and therapies of the future.

## Notes

### Competing Interest Statement

The authors have declared no competing interest.

### Author Declarations

All data have been obtained from open access sources such as cBioportal, Geneminer, Oncomine and KMPlot.

## References

1. Sporikova, Z., et al., Genetic Markers in Triple-Negative Breast Cancer. Clinical Breast Cancer, 2018. 18(5): p. e841–e850.

2. Sung, H., et al., Global cancer statistics 2020: GLOBOCAN estimates of incidence and mortality worldwide for 36 cancers in 185 countries. CA: A Cancer Journal for Clinicians, 2021. n/a(n/a).

3. Coughlin, S.S., Epidemiology of Breast Cancer in Women. Adv Exp Med Biol, 2019. 1152: p. 9–29.

4. Russnes, H.G., et al., Breast Cancer Molecular Stratification: From Intrinsic Subtypes to Integrative Clusters. The American Journal of Pathology, 2017. 187(10): p. 2152–2162.

5. Han, Y. and X. Wang, The emerging roles of KPNA2 in cancer. Life Sciences, 2020. 241: p. 117140.

6. Li, J., et al., KPNA2 promotes metabolic reprogramming in glioblastomas by regulation of c-myc. Journal of Experimental & Clinical Cancer Research, 2018. 37(1): p. 194.

7. Zhang, Y., et al., Karyopherin alpha 2 is a novel prognostic marker and a potential therapeutic target for colon cancer. Journal of experimental & clinical cancer research: CR, 2015. 34: p. 145–145.

8. Gao, C.-L., et al., Karyopherin subunit-α 2 expression accelerates cell cycle progression by upregulating CCNB2 and CDK1 in hepatocellular carcinoma. Oncology letters, 2018. 15(3): p. 2815–2820.

9. Huang, L., et al., KPNA2 promotes migration and invasion in epithelial ovarian cancer cells by inducing epithelial-mesenchymal transition via Akt/GSK-3β/Snail activation. Journal of Cancer, 2018. 9(1): p. 157–165.

10. Ma, A., et al., USP1 inhibition destabilizes KPNA2 and suppresses breast cancer metastasis. Oncogene, 2019. 38(13): p. 2405–2419.

11. Noetzel, E., et al., Nuclear transport receptor karyopherin-α2 promotes malignant breast cancer phenotypes in vitro. Oncogene, 2012. 31(16): p. 2101–2114.

12. Alshareeda, A.T., et al., KPNA2 is a nuclear export protein that contributes to aberrant localisation of key proteins and poor prognosis of breast cancer. British journal of cancer, 2015. 112(12): p. 1929–1937.

13. Pavlou, M.P., et al., Integrating Meta-Analysis of Microarray Data and Targeted Proteomics for Biomarker Identification: Application in Breast Cancer. Journal of Proteome Research, 2014. 13(6): p. 2897–2909.

14. Huang, L., et al., KPNA2 promotes cell proliferation and tumorigenicity in epithelial ovarian carcinoma through upregulation of c-Myc and downregulation of FOXO3a. Cell Death & Disease, 2013. 4(8): p. e745–e745.

15. Gao, J., et al., Integrative analysis of complex cancer genomics and clinical profiles using the cBioPortal. Sci Signal, 2013. 6(269): p. pl1.

16. Mizuno, H., et al., PrognoScan: a new database for meta-analysis of the prognostic value of genes. BMC Medical Genomics, 2009. 2(1): p. 18.

17. Mizuno, H., et al., PrognoScan: a new database for meta-analysis of the prognostic value of genes. BMC medical genomics, 2009. 2: p. 18–18.

18. Györffy, B., et al., An online survival analysis tool to rapidly assess the effect of 22,277 genes on breast cancer prognosis using microarray data of 1,809 patients. Breast Cancer Res Treat, 2010. 123(3): p. 725–31.

19. Thomssen, C., et al., St. Gallen/Vienna 2021: A Brief Summary of the Consensus Discussion on Customizing Therapies for Women with Early Breast Cancer. Breast Care, 2021. 16(2): p. 135–143.

20. Jézéquel, P., et al., bc-GenExMiner 4.5: new mining module computes breast cancer differential gene expression analyses. Database (Oxford), 2021. 2021.

21. Jézéquel, P., et al., bc-GenExMiner: an easy-to-use online platform for gene prognostic analyses in breast cancer. Breast Cancer Res Treat, 2012. 131(3): p. 765–75.

22. Parry, M., Introducing the Metastatic Breast Cancer Project: a novel patient-partnered initiative to accelerate understanding of MBC. ESMO open, 2018. 3(7): p. e000452–e000452.

23. Lefebvre, C., et al., Mutational Profile of Metastatic Breast Cancers: A Retrospective Analysis. PLoS medicine, 2016. 13(12): p. e1002201–e1002201.

24. Curtis, C., et al., The genomic and transcriptomic architecture of 2,000 breast tumours reveals novel subgroups. Nature, 2012. 486(7403): p. 346–52.

25. Rueda, O.M., et al., Dynamics of breast-cancer relapse reveal late-recurring ER-positive genomic subgroups. Nature, 2019. 567(7748): p. 399–404.

26. Pereira, B., et al., The somatic mutation profiles of 2,433 breast cancers refines their genomic and transcriptomic landscapes. Nat Commun, 2016. 7: p. 11479.

27. Berger, A.C., et al., A Comprehensive Pan-Cancer Molecular Study of Gynecologic and Breast Cancers. Cancer Cell, 2018. 33(4): p. 690-705.e9.

28. Richardson, A.L., et al., X chromosomal abnormalities in basal-like human breast cancer. Cancer Cell, 2006. 9(2): p. 121–132.

29. Clare, S.E. and P.L. Shaw, “Big Data” for breast cancer: where to look and what you will find. npj Breast Cancer, 2016. 2(1): p. 16031.

30. Mehmood, R., et al., Molecular profiling of nucleocytoplasmic transport factor genes in breast cancer. Heliyon, 2021. 7(1): p. e06039.

31. Tovar, E.A. and C.R. Graveel, MET in human cancer: germline and somatic mutations. Annals of Translational Medicine, 2017. 5(10): p. 205–205.

32. Zhang, X., R. Zhang, and J. Yu, New Understanding of the Relevant Role of LINE-1 Retrotransposition in Human Disease and Immune Modulation. Frontiers in cell and developmental biology, 2020. 8: p. 657–657.

33. Wu, X., et al., The human oncogene SCL/TAL1 interrupting locus (STIL) promotes tumor growth through MAPK/ERK, PI3K/Akt and AMPK pathways in prostate cancer. Gene, 2019. 686: p. 220–227.

34. Cui, X., et al., Increased expression of KPNA2 predicts unfavorable prognosis in ovarian cancer patients, possibly by targeting KIF4A signaling. Journal of Ovarian Research, 2021. 14(1).

35. Wang, J., et al., KIF2A silencing inhibits the proliferation and migration of breast cancer cells and correlates with unfavorable prognosis in breast cancer. BMC Cancer, 2014. 14(1): p. 461.

36. Tang, J., et al., Genome-wide expression profiling of glioblastoma using a large combined cohort. Sci Rep, 2018. 8(1): p. 15104–12.

37. Li, H.N., et al., ZW10 interacting kinetochore protein may serve as a prognostic biomarker for human breast cancer: An integrated bioinformatics analysis. Oncology Letters, 2020.

38. Chen, M., et al., Structure and regulation of human epithelial cell transforming 2 protein. Proceedings of the National Academy of Sciences, 2020. 117(2): p. 1027–1035.

39. Alfarsi, L.H., et al., CDC20 expression in oestrogen receptor positive breast cancer predicts poor prognosis and lack of response to endocrine therapy. Breast Cancer Res Treat, 2019. 178(3): p. 535–544.

40. Gluz, O., et al., Nuclear karyopherin α2 expression predicts poor survival in patients with advanced breast cancer irrespective of treatment intensity. International Journal of Cancer, 2008. 123(6): p. 1433–1438.

41. Dankof, A., et al., KPNA2 protein expression in invasive breast carcinoma and matched peritumoral ductal carcinoma in situ. Virchows Arch, 2007. 451(5): p. 877–881.

42. Dahl, E., et al., Molecular Profiling of Laser-Microdissected Matched Tumor and Normal Breast Tissue Identifies Karyopherin α2 as a Potential Novel Prognostic Marker in Breast Cancer. Clinical Cancer Research, 2006. 12(13): p. 3950–3960.

