## Supplemental data for "The role of *KPNA2* mutations in breast cancer prognosis: A survey of publicly available databases"

**Supplementary information**

Table S 1 Corresponding statistical tests and study attributes reported for analyses conducted on KPNA2 expression levels in breast cancer studies selected from cBioportal

| Clinical Attribute | Attribute | Statistical Test | p-Value | q-Value |
| --- | --- | --- | --- | --- |
| Integrative Cluster | Patient | Chi-squared Test | 0 | 0 |
| PAM50 + Claudin-low subtype | Patient | Chi-squared Test | 3.44E-15 | 3.65E-13 |
| 3-Gene classifier subtype | Patient | Chi-squared Test | 8.68E-13 | 6.14E-11 |
| HER2 status measured by SNP6 | Patient | Chi-squared Test | 6.04E-11 | 3.20E-09 |
| Lymph nodes examined positive | Patient | Chi-squared Test | 4.67E-06 | 1.98E-04 |
| Neoplasm Histologic Grade | Sample | Chi-squared Test | 2.60E-05 | 9.20E-04 |
| ER Status By IHC | Sample | Chi-squared Test | 4.19E-05 | 1.27E-03 |
| ER positivity scale other | Sample | Chi-squared Test | 6.49E-05 | 1.57E-03 |
| Nottingham prognostic index | Patient | Wilcoxon Test | 6.65E-05 | 1.57E-03 |
| PR positivity scale other | Sample | Chi-squared Test | 4.84E-04 | 9.77E-03 |
| Number of Samples Per Patient | Patient | Chi-squared Test | 5.07E-04 | 9.77E-03 |
| Cohort | Patient | Chi-squared Test | 7.62E-04 | 0.0135 |
| HER2 IHC score | Sample | Wilcoxon Test | 9.28E-04 | 0.015 |
| HER2 Status | Sample | Chi-squared Test | 1.03E-03 | 0.015 |
| Mutation Count | Sample | Wilcoxon Test | 1.06E-03 | 0.015 |
| Fraction Genome Altered | Sample | Wilcoxon Test | 1.85E-03 | 0.0245 |
| HER2 FISH status | Sample | Chi-squared Test | 2.25E-03 | 0.028 |
| Diagnosis Age | Patient | Wilcoxon Test | 3.49E-03 | 0.0411 |
| ER Status IHC Percent Positive | Sample | Chi-squared Test | 4.01E-03 | 0.0447 |

This table summarizes the statistical tests used across the studies reported in the main text (cBioportal), representing the clinicopathologic parameters that are found to differ significantly between in the presence and absence of alterations in KPNA2 levels.

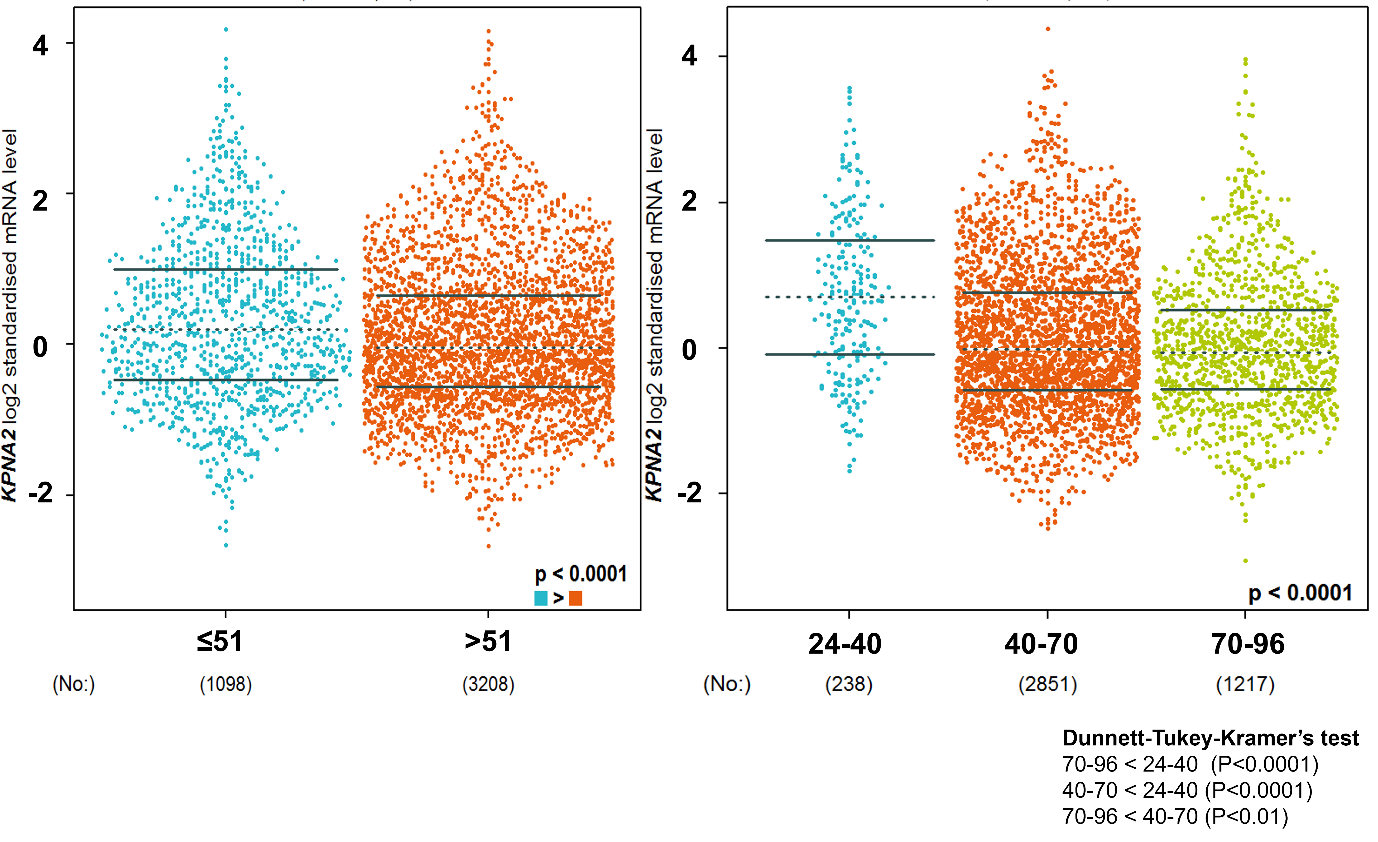

Fig S 1 Beeswarm plots of KPNA2 mRNA expression levels as a function of patient age.

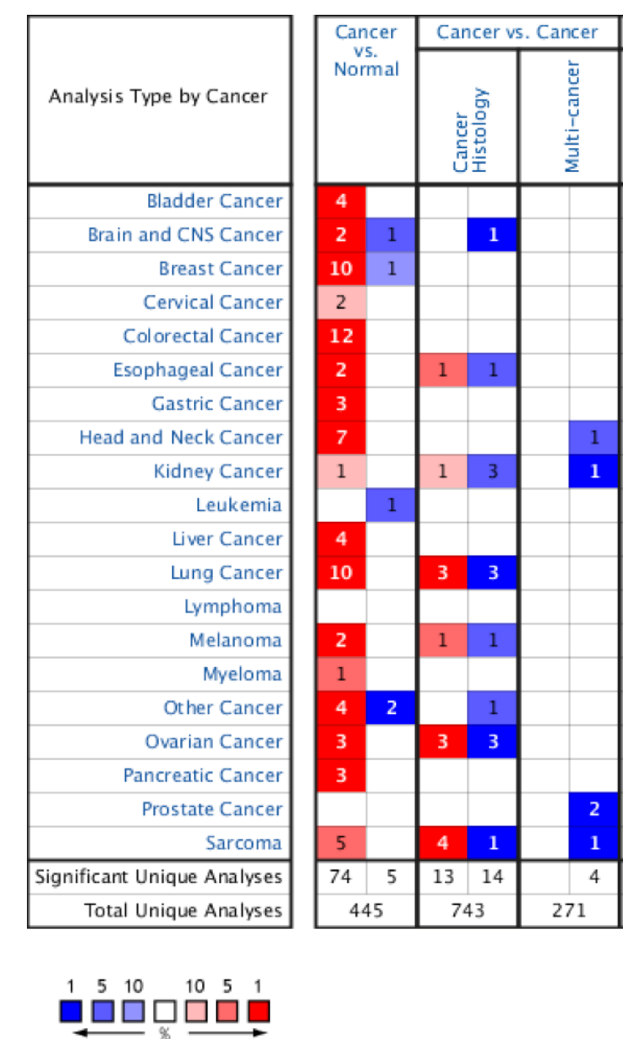

Fig S 2 Pan cancer evaluation of KPNA2 transcriptional levels in different cancer types. Blue represents lower levels of expression, while red indicates overexpression of KPNA2. The numbers presented represent the number of studies meeting the search criteria for downstream analysis.

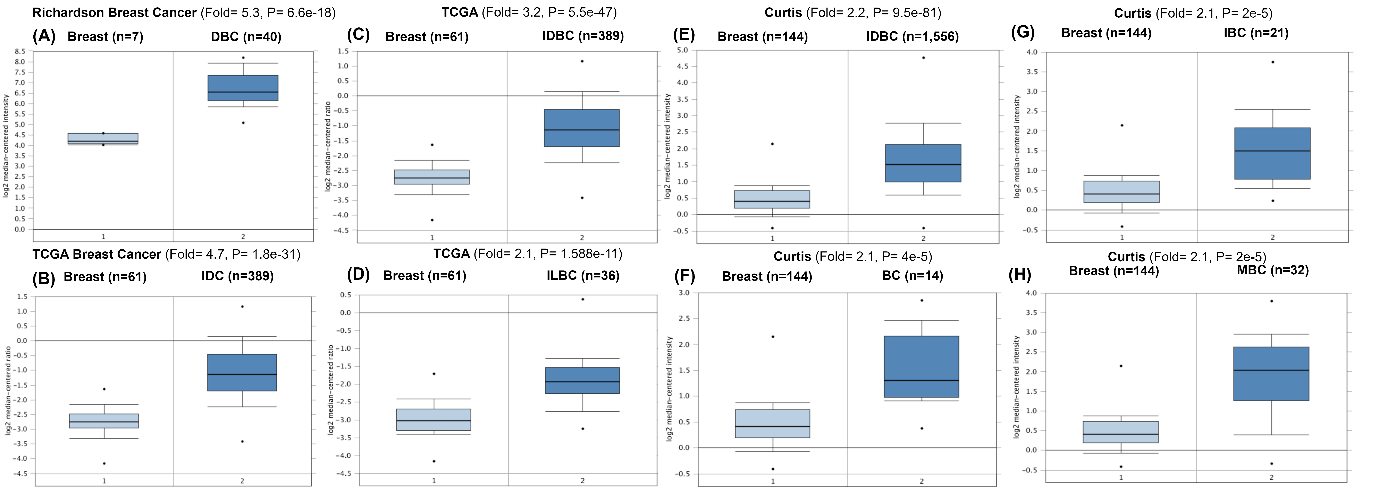

Fig S 3 Corresponding boxplots of KPNA2 mRNA expression profiles in different breast cancer types (left boxes- 1) versus normal breast tissue (right boxes- 2) as obtained from Oncomine. Data presented for ductal breast carcinoma (DBC) **(A)**, invasive ductal carcinoma (IDC) **(B)**, invasive ductal breast carcinoma (IDBC) **(C)**, invasive lobal breast carcinoma (ILBC) **(D)**, invasive ductal breast carcinoma (IDBC) **(E),** breast carcinoma (BC) **(F),** invasive breast carcinoma (IBC) **(G)** and medullary breast carcinoma (MBC) **(H)**.

**The prognostic value of KPNA2 in breast cancer across intrinsic subtypes**

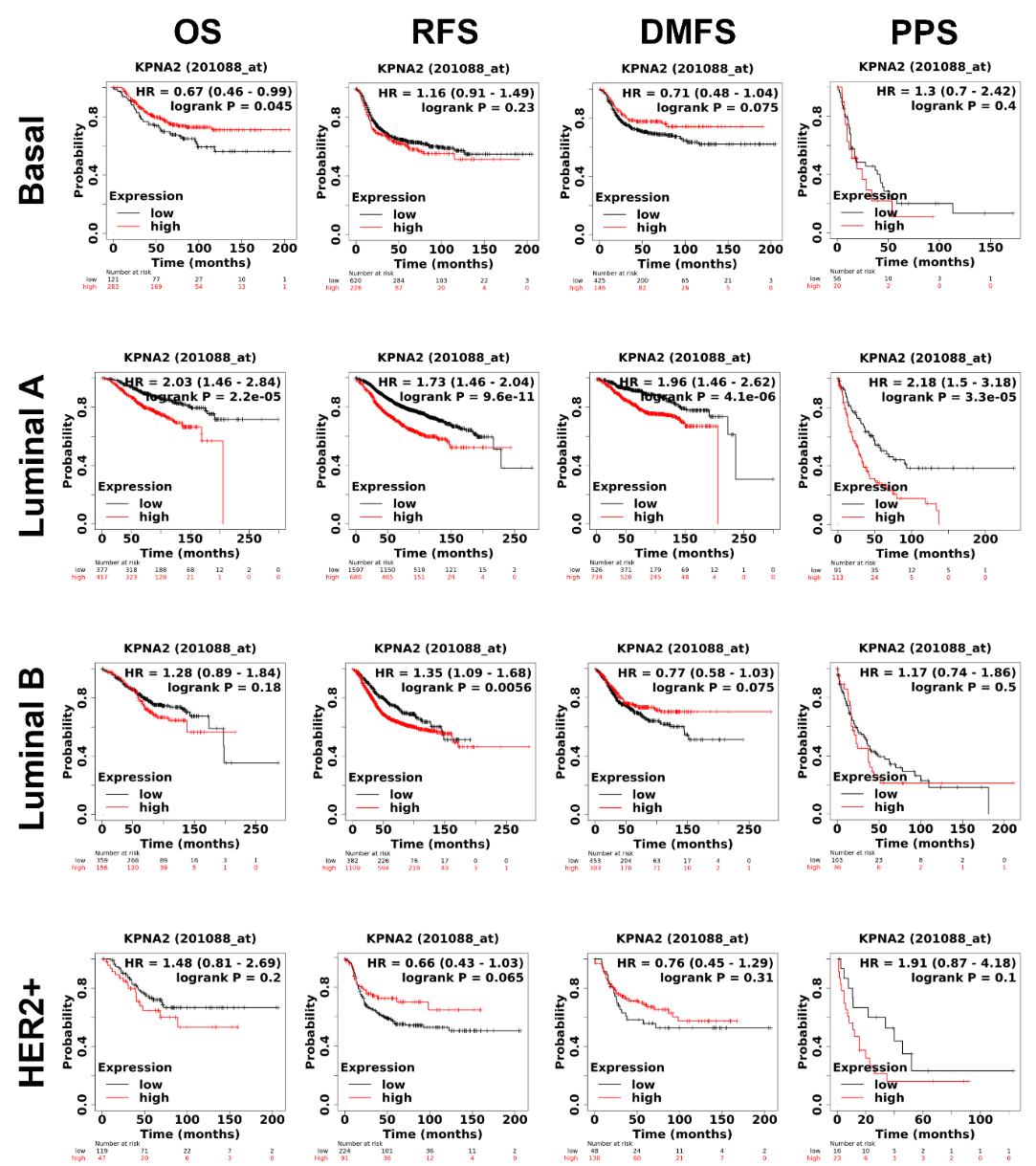

Fig S 4 Kaplan Meier plots of breast cancer patient survival endpoints as a function of KPNA2 expression levels across intrinsic molecular subtypes. **OS:** Overall Survival, **RFS:** Relapse-Free Survival, **DMFS:** Disease-Metastatic Free Progression Survival, **PPS:** Post-Progression Survival.

Corresponding false discovery rates for Kaplan-Meier plots in Fig S 4 are presented in Table S 2:

Table S 2 Breast cancer subtype expected False Discovery Rate (FDR) per prognostic endpoint from KM plotter

| **Breast cancer** | **OS** | **RFS** | **DMFS** | **PPS** |
| --- | --- | --- | --- | --- |
| All | 1% | 1% | 1% | 1% |
| Basal | >50% | 100% | 100% | 100% |
| Luminal A | 1% | 1% | 1% | 1% |
| Luminal B | 100% | >50% | 100% | 100% |
| HER-2 | 100% | 100% | 100% | 100% |

**Co-expression patterns of KPNA2 and other genes according to hormone receptor status**

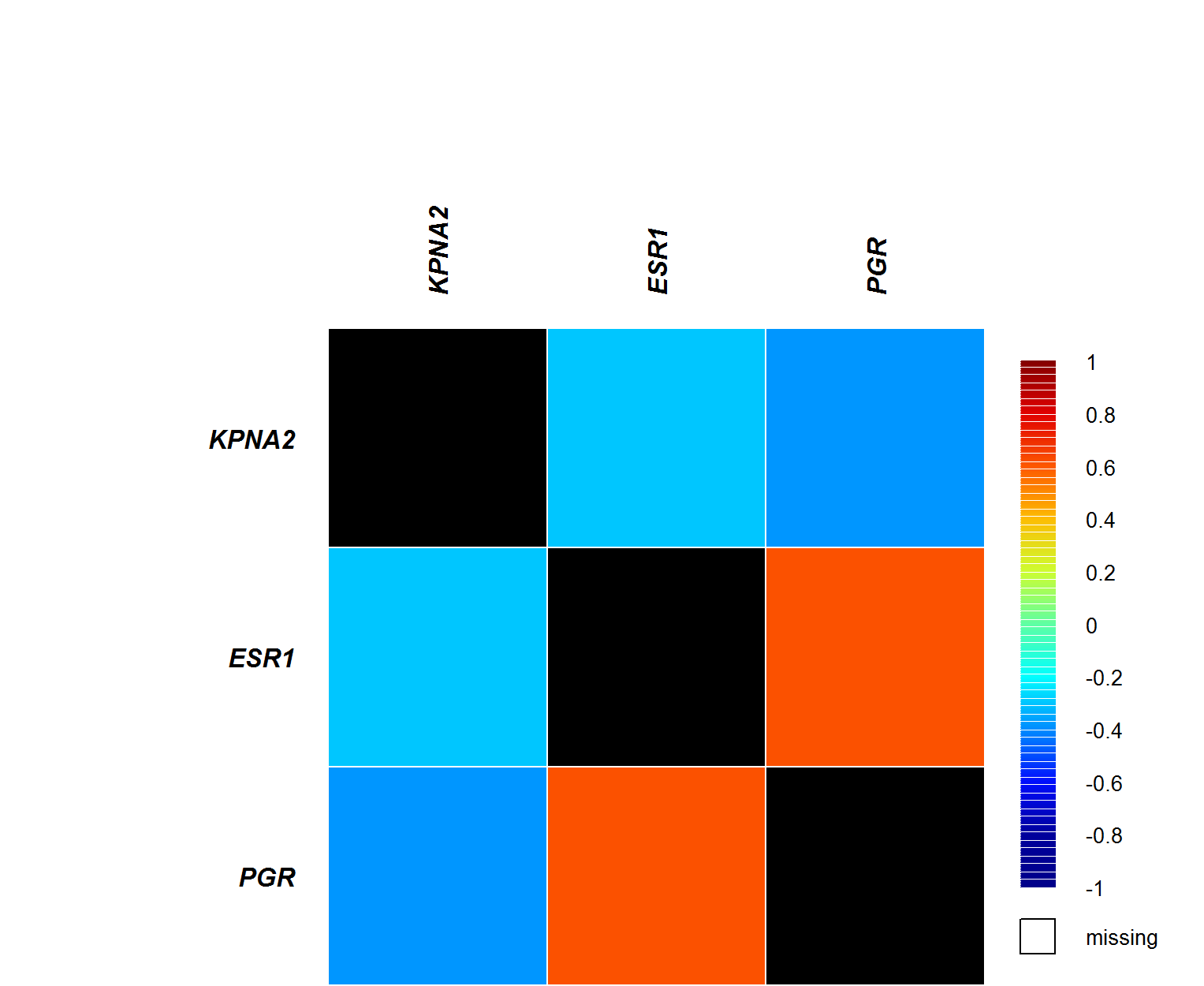

Fig S 5 Heatmap of KPNA2 mRNA co-expression with estrogen receptor (ESR1) and progesterone receptors (PGR) across all breast cancers, obtained from Geneminer.

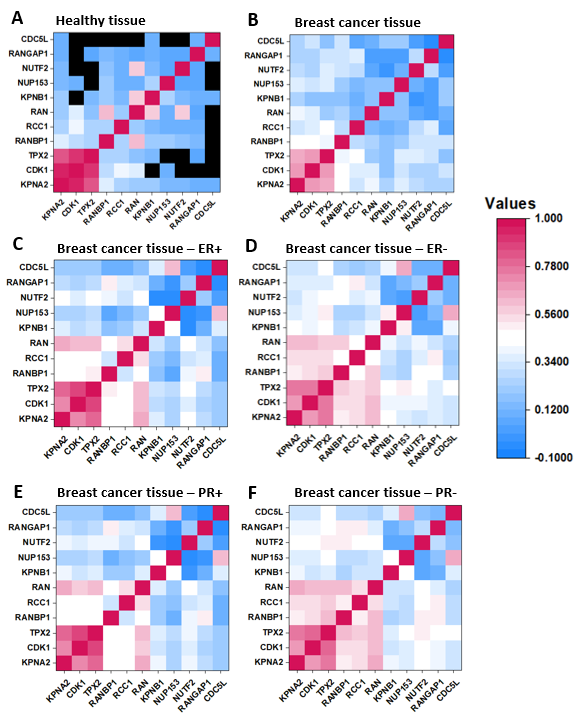

Fig S 6 **Differences between KPNA2-correlated genes in healthy and a variety of breast cancer tissues.** The colour shade indicates the level of confidence that two proteins are functionally associated, given the overall expression data. Co-expression scores are based on mRNA expression levels and protein co-regulation. The black colour indicates the absence of data. Heatmaps of protein-protein correlation matrices in healthy tissue **(A)**, breast cancer tissue **(B)**, ER+ (**C)**, ER- (**D)**, PR+ **(E)**, and PR- breast cancer tissues.

Table S 3 Categorizing multiple KPNA2 co-expressed genes according to their biological role and cancer implications obtained from Kyoto Encyclopedia of Genes and Genomes (KEGG) <https://doi.org/10.1093/nar/28.1.27.> In association with [UniProt and the Human Proteome Atlas,](https://www.uniprot.org/) this resource provides a link of gene functions to published literature. Gene subcellular location obtained from EMBL-EBI GO Annotations ([QuickGO (ebi.ac.uk)](https://www.ebi.ac.uk/QuickGO/)).

| **Co-expressed gene** | **Gene name** | **Subcellular location** | **Biological role** | **Cancer implications** |
| --- | --- | --- | --- | --- |
| ***Cell growth*** | | | | |
| CCNA2 | Cyclin A2 | Nucleoplasm, cytosol | Signal transduction of DNA replication proteins and initiation factors | Transcription dysregulation |
| ANLN | Actin-binding protein anillin | Nucleoplasm, midbody | Microfilament binding to form protein cytoskeleton maintaining the cell structure | Promotes cytokine secretion inducing metastasis |
| STIL | SCL-interrupting locus protein | Developmental protein | Regulates protein duplication and replication | Mutated STIL impairs cell proliferation and development |
| ***DNA synthesis / replication*** | | | | |
| TOP2A | DNA topoisomerase 2-alpha | Nucleoplasm, Nucleoli | Chromosome condensation and DNA replication | induces platinum drug resistance |
| WHSC1 | Wolf-Hirschhorn syndrome candidate 1 |  | Involved in amino acid metabolism | Promotes cancer cell migration and invasion by activating mTOR signalling causing transcription mis regulation. |
| NUF2 | Kinetochore protein Nuf2 | Nucleoplasm, kinetochore, cytosol | Chromatin formation proteins  Defective genetic information processing | |
| MAD2L1 | Mitotic arrest deficient 2 like 1 | Nucleoplasm |  |  |
| **DNA repair / ubiquitination** | | | | |
| FAM83D | Family with sequence similarity 83 member D |  | DNA replication, cross linking, and double strand repair  Abnormal chromosomal accumulation | |
| UBE2T | ubiquitin-conjugating enzyme E2 T | Nucleoplasm, Nucleoli |  |  |
| BUB1B | Mitotic checkpoint serine/threonine-protein kinase BUB1 beta | Cytosol | Nucleotide metabolism and P53 signalling  Impaired DNA repair mechanisms and biosynthesis | |
| RRM2 | Ribonucleoside-diphosphate reductase subunit matric protein 2 | Cytosol |  |  |
| TACC3 | transforming acidic coiled-coil-containing protein 3 |  | Messenger RNA biogenesis | |
| MYBL2 | Myb-related protein B |  | Transcription factor regulating DNA repair  Direct activation of anti-apoptotic genes, such as clusterin | |
| DTL | D-dopachrome tautomerase like | Nucleoplasm, Nucleoli, Cytosol | Protein ubiquitination and ensures cell integrity during its division  Promotes translesion DNA synthesis | |
| UHRF1 | Ubiquitin-protein ligase |  | Maintains DNA methylation in cells  Prevents DNA damage response and apoptosis | |
| DSCC1 | DNA Replication and Sister Chromatid Cohesion 1 |  | DNA replication  Alters DNA repair processes | |
| TRIP13 | Pachytene checkpoint protein 2 | cytoplasm | Involved in chromosome structure development  Alters upstream DNA repair processes | |
| ***Cell division*** | | | | |
| MELK | Maternal embryonic leucine zipper kinase | Cytoplasmic, nuclear | Transferring phosphorous containing groups for metabolism | Oncogene that drives mitosis progression |
| NEK2 | NIMA related kinase 2 | Nucleoplasm, centrosome | Cell signal processes and protein cytoskeleton formation  Induces cytokinesis involved in tubulin binding | |
| KIF2C | Kinesin family member 2C | Cytoplasm |  |  |
| KIF14 | Kinesin family member 14 | Midbody ring, cytosol | Mediates localisation of spindle proteins prior to mitosis | Aberrant spindle protein formation |
| CDCA8 | Cell division cycle associated 8 | Nucleoli | Required for chromosomal stability of the mitotic spindle | Induces chromosomal alignment by forming chromatin proteins |
| CDK1 | Cyclin-dependent kinase 1 | Nucleoplasm, cytosol | Metabolism and phosphorylation of DNA replication factors | Cellular senescence and promotes mitosis |
| CDKN3 | Cyclin dependent kinase inhibitor 3 | Cytosol | Protein phosphatases involved in cellular metabolism | Alters immune response ability to regulate tumour cell cycle |
| NUSAP1 | Nucleolar and spindle associated protein 1 | Nucleoplasm, nucleoli fibrillar centre | microtubule-associated protein | Affects mitotic spindle organization |
| ZWINT | ZW10 interacting kinetochore protein | Nucleoplasm, nuclear bodies, cytosol | Membrane trafficking of mitotic checkpoint proteins  Aberrant protein membrane trafficking | |
| ECT2 | Epithelial cell transforming 2 | Nucleoplasm, cytosol |  |  |
| CEP55 | Centrosomal protein CEP55 | Plasma membrane, Midbody, centriolar satellite |  |  |
| CASC5 | Cancer susceptibility candidate 5 | Nucleoplasm, nuclear bodies | Mitotic spindle assembly | Abnormal mitosis checkpoint signalling |
| CCNB2 | G2/mitotic-specific cyclin-B2 | Retrieving data. Wait a few seconds and try to cut or copy again. | P53 signalling pathway | Induces cellular senescence |
| CDC45 | Cell division control protein 45 | Nucleoplasm, centrosome, cytosol | Maintains chromosomal DNA replication | Drives aberrant cell division by increasing DNA replication proteins |
| CDC20 | Cell division cycle 20 | Nucleoplasm, cytosol | Cofactor of APC complex |  |
| HMMR | Hyaluronan-mediated motility receptor | cytoplasm | A mitotic spindle-binding protein | Promotes cell mobility by activating MAP kinase pathway |
| MLF1IP | centromere protein U | Nucleus, cytoplasm | Chromatin formation protein | Alters genetic information processing |
| KIAA0101 | clamp associated factor | Nucleus, cytoplasm | Regulation of centrosomes | Genomic instability due to centrosome disruption |
| ZNF367 | zinc finger protein 367 | Nucleus, cytoplasm | Regulates DNA-binding transcription factors | Promotes metastasis through HIPPO pathway |
| NUSAP1 | Nucleolar and spindle associated protein 1 | Nucleoplasm, nucleoli fibrillar centre | Mitotic phosphoprotein | Destabilises spindle microtubule |
| TPX2 | Targeting protein for Xklp2 | Nucleoplasm, cytokinetic bridge, mitotic spindle | Microtubule organisation | Abnormal mitotic spindles assembly |
